# Vaccine effectiveness against SARS-CoV-2 transmission to household contacts during dominance of Delta variant (B.1.617.2), August-September 2021, the Netherlands

**DOI:** 10.1101/2021.10.14.21264959

**Authors:** Brechje de Gier, Stijn Andeweg, Jantien A. Backer, RIVM COVID-19 surveillance and epidemiology team, Susan J.M. Hahné, Susan van den Hof, Hester E. de Melker, Mirjam J. Knol

## Abstract

We estimated vaccine effectiveness against onward transmission by comparing secondary attack rates among household members between vaccinated and unvaccinated index cases, based on source and contact tracing data collected when Delta variant was dominant. Effectiveness of full vaccination of the index against transmission to fully vaccinated household contacts was 40% (95% confidence interval (CI) 20-54%), which is in addition to the direct protection of vaccination of contacts against infection. Effectiveness of full vaccination of the index against transmission to unvaccinated household contacts was 63% (95%CI 46-75%). We previously reported effectiveness of 73% (95%CI 65-79%) against transmission to unvaccinated household contacts for the Alpha variant.

---

Early August 2021 we reported vaccine effectiveness against SARS-CoV-2 transmission and infections among household and other close contacts of confirmed cases [1]. This study was based on source and contact tracing data collected in February-May 2021, during which time the wildtype and Alpha variant of SARS-CoV-2 were dominating. From May 29 to July 4, a transition took place where the Delta variant took over from Alpha and became dominant, with over 85% of sequenced isolates from week of July 5-July 11 pertaining the Delta variant. At the end of June, several non-pharmaceutical measures were relaxed in the Netherlands [2]. This relaxation, combined with the co-occurring emergence of Delta, was followed by a large increase of SARS-CoV-2 infections (Figure 1). This increase was mainly driven by infections in young unvaccinated people.

**Fig. 1.**
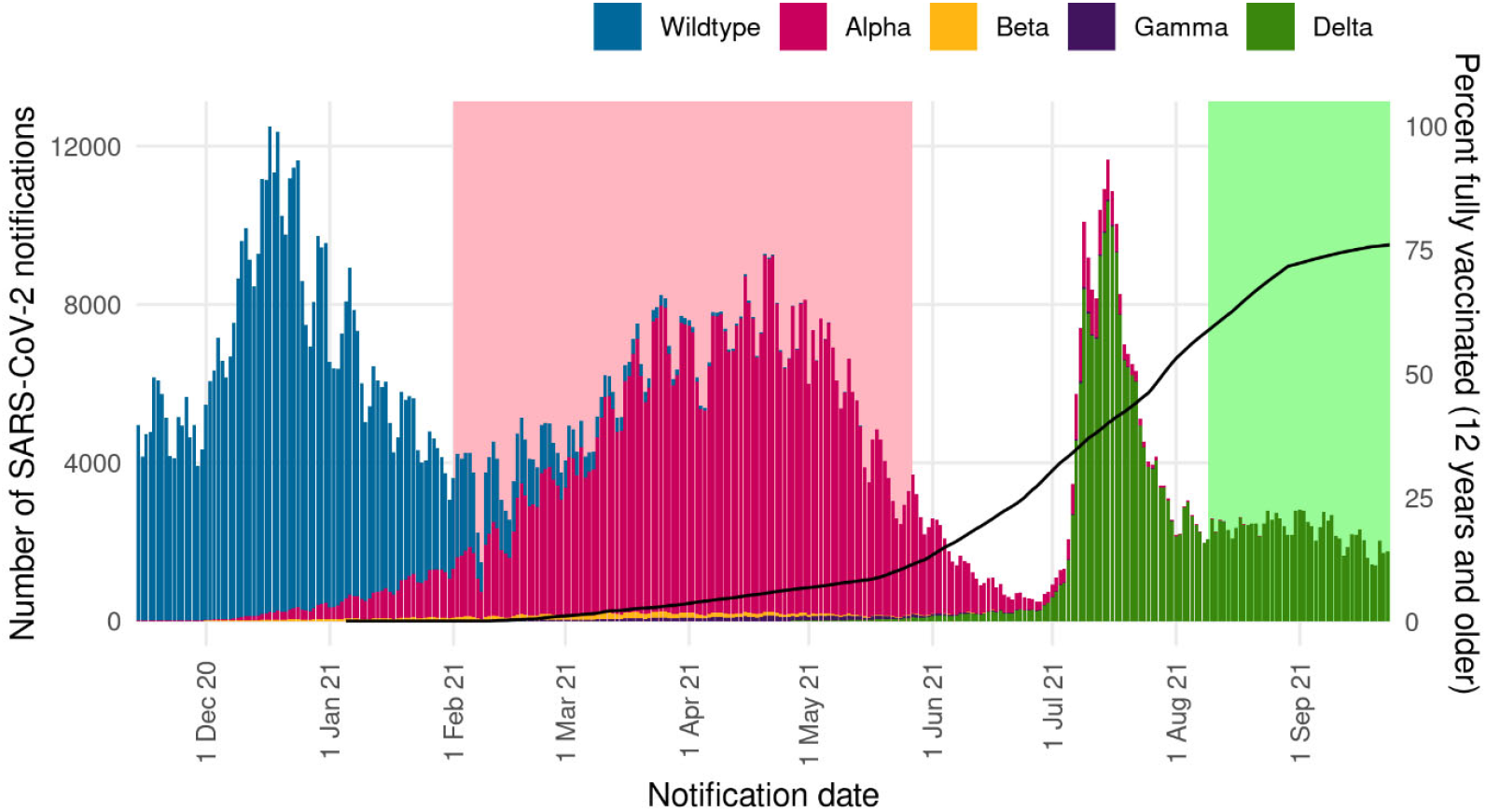
Number of cases per day in the Netherlands by symptom onset, colored by SARS-CoV-2 variant, and percent of the Dutch population fully vaccinated (defined as 14 or more days post-second-dose or 28 or more days post-single dose (Janssen), black line). The light green area indicates the study period of the study presented here. The pink area indicates the study period of our previous report. The share of SARS-CoV-2 variant results from fitting exponential growth curves to weekly surveillance data of sequenced samples [3].

A result of this large and fast increase in the number of cases was that Municipal Health Services (MHS) lacked capacity to perform full source and contact tracing in July and the beginning of August. Full-scale source and contact tracing has been resumed by all MHS since August 9, 2021. Therefore, our analysis of vaccine effectiveness against transmission (VET) of the Delta variant is only possible for data collected since August 9, 2021. We end our study period at September 24, 2021, as since September 25 unvaccinated people require a negative test or proof of recovery to enter bars, restaurants and events, which will impact testing behavior differentially by vaccination status [4]. During the study period, over 97% of sequenced isolates pertained Delta [3].

Until July 2021, all household contacts of confirmed cases had to quarantine for 10 days, and were urged to get tested on day 1 and day 5 after exposure and in case of symptoms. If a contact was tested negative on day 5, they could end the quarantine. On July 8, 2021, a policy change was implemented, and fully vaccinated household contacts of confirmed cases no longer had to quarantine. These fully vaccinated contacts were still strongly advised to get tested on day 5 and to practice social distancing until day 10.

A full description of the data and methods used can be found in our previous report [1]. In short, the VE against transmission (VET) was estimated by comparing the secondary attack rate (SAR) among household contacts of confirmed index cases by vaccination status of the index case: 1 – (SAR _vaccinated index_ / SAR _unvaccinated index_) * 100%. An index case is a person with a positive SARS-CoV-2 test, who according to the source tracing interview, was most likely not infected at home. Index cases and household contacts aged 12 years or older are included in the analysis, as all Dutch inhabitants above the age of 12 years are eligible for vaccination since July 2021. Partly vaccinated was defined as having received the first dose of a 2-dose schedule with a time since vaccination of at least 14 days. Fully vaccinated was defined as having completed a 2-dose schedule with a time since vaccination of at least 14 days, or the 1-dose Janssen schedule with a time since vaccination of at least 28 days.

VET was estimated using a binomial generalized linear model. For parameter fitting we used the generalized estimating equations (GEE) approach with exchangeable correlation structure to account for clustering of contacts belonging to the same index case. All models included age group of the index and contact (12-17, 18-29, 30-49, 50-74 and 75+ years), vaccination status of the contact (not, partly or fully vaccinated) and week of notification date of the index case as covariates.

The final dataset contained 7,771 contacts of 4,921 index cases. Of the index cases, 2,602 (33.5%) were fully vaccinated and 912 (11.7%) were partly vaccinated. Of contacts, 4,189 (53.9%) were fully vaccinated and 641 were partly vaccinated (8.2%). The predominance of unvaccinated index cases is the result of vaccine effectiveness against infection, as 71% of all adults was fully vaccinated at the start of the study period [5]. Characteristics of indexes and contacts are shown in Table 1.

**Table 1.** Characteristics of index cases, by vaccination status of the index and characteristics of contacts, by vaccination status of the contact. NA = not applicable.

|  | Index cases |  |  | Household contacts |  |  |
| --- | --- | --- | --- | --- | --- | --- |
|  | Unvaccinated<br>(column %) | Partly vaccinated<br>(column %) | Fully vaccinated<br>(column %) | Unvaccinated<br>(column %) | Partly vaccinated<br>(column %) | Fully vaccinated<br>(column %) |
| Total | 2641 | 540 | 1740 | 2941 | 641 | 4189 |
| Gender |  |  |  |  |  |  |
| Female | 1480 (56%) | 280 (52%) | 871 (50%) | 1517 (52%) | 313 (49%) | 2106 (50%) |
| Male | 1161 (44%) | 260 (48%) | 869 (50%) | 1379 (47%) | 320 (50%) | 2026 (48%) |
| Unknown/ other | 0 (0%) | 0 (0%) | 0 (0%) | 45 (2%) | 8 (1%) | 57 (1%) |
| Age group |  |  |  |  |  |  |
| 12-17 | 1005 (38%) | 174 (32%) | 45 (3%) | 903 (31%) | 172 (27%) | 127 (3%) |
| 18-29 | 823 (31%) | 229 (42%) | 549 (32%) | 718 (24%) | 216 (34%) | 673 (16%) |
| 30-49 | 616 (23%) | 101 (19%) | 438 (25%) | 910 (31%) | 176 (27%) | 1460 (35%) |
| 50-74 | 183 (7%) | 33 (6%) | 631 (36%) | 383 (13%) | 77 (12%) | 1841 (44%) |
| 75+ | 14 (1%) | 3 (1%) | 77 (4%) | 27 (1%) | 0 (0%) | 88 (2%) |
| Vaccine received |  |  |  |  |  |  |
| Comirnaty | NA | 483 (89%) | 963 (55%) | NA | 505 (79%) | 2544 (61%) |
| Spikevax | NA | 43 (8%) | 87 (5%) | NA | 53 (8%) | 389 (9%) |
| Vaxzevria | NA | 14 (3%) | 273 (16%) | NA | 4 (1%) | 350 (8%) |
| Janssen | NA | NA | 417 (24%) | NA | NA | 420 (10%) |
| Unknown | NA | 0 (0%) | 0 (0%) | NA | 78 (12%) | 486 (12%) |
| Household composition |  |  |  |  |  |  |
| Two adults without children | 825 (31%) | 165 (31%) | 977 (56%) | 634 (22%) | 160 (25%) | 1173 (28%) |
| Two adults with child(ren) | 723 (27%) | 153 (28%) | 273 (16%) | 900 (31%) | 184 (29%) | 1041 (25%) |
| Single adult with child(ren) | 573 (22%) | 78 (14%) | 83 (5%) | 496 (17%) | 79 (12%) | 309 (7%) |
| Other | 520 (20%) | 144 (27%) | 407 (23%) | 911 (31%) | 218 (34%) | 1666 (40%) |
| Month of notification of index case |  |  |  |  |  |  |
| aug | 1410 (53%) | 429 (79%) | 916 (53%) | 1542 (52%) | 486 (76%) | 2178 (52%) |
| sep | 1231 (47%) | 111 (21%) | 824 (47%) | 1399 (48%) | 155 (24%) | 2011 (48%) |

Vaccination status by age reflects the roll-out of vaccination from old to young. Table 2 shows the vaccination status of contacts by vaccination status of index cases. Of unvaccinated index cases, 59.1% of household contacts were unvaccinated as well, while only 11.6% of household contacts of vaccinated index cases were unvaccinated.

**Table 2.** Vaccination status of contacts relative to vaccination status of index cases

| Vaccination status contact | Unvaccinated index, (column %) | Partly vaccinated index, (column %) | Fully vaccinated index, (column %) |
| --- | --- | --- | --- |
| Unvaccinated | 2517 (59.1) | 121 (13.3) | 303 (11.6) |
| Partly vaccinated | 235 (5.5) | 177 (19.4) | 229 (8.8) |
| Fully vaccinated | 1505 (35.4) | 614 (67.3) | 2070 (79.6) |

Table 3 shows a lower crude SAR among unvaccinated household contacts for vaccinated index cases compared to unvaccinated index cases (13% vs. 22%) and a corresponding adjusted vaccine effectiveness of 63% (95%CI 46-75%) against transmission. Among fully vaccinated household contacts, the crude SAR was similar for fully vaccinated index cases compared to unvaccinated index cases (11% vs. 12%), but this was confounded by age of the index – both SAR and proportion of vaccinated index cases are higher in the oldest age groups (Supplementary Table S1). After adjustment, the effectiveness of full vaccination of the index case was 40% (95%CI 20-54%).

**Table 3.**
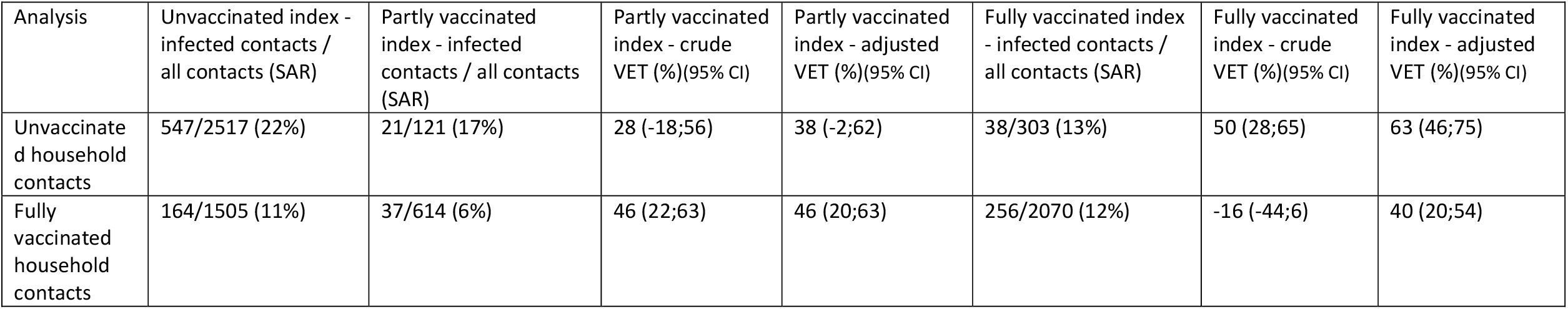
Secondary attack rate of SARS-CoV-2 infection by vaccination status of the index case (≥ 12 years), crude vaccine effectiveness against transmission (VET) and VET adjusted for age group of the index case and contact and week of notification date of the index case.

| Analysis | Unvaccinated index - infected contacts / all contacts (SAR) | Partly vaccinated index - infected contacts / all contacts (SAR) | Partly vaccinated index - crude VET (%) (95% CI) | Partly vaccinated index - adjusted VET (%) (95% CI) | Fully vaccinated index - infected contacts / all contacts (SAR) | Fully vaccinated index - crude VET (%) (95% CI) | Fully vaccinated index - adjusted VET (%) (95% CI) |
| --- | --- | --- | --- | --- | --- | --- | --- |
| Unvaccinated household contacts | 547/2517 (22%) | 21/121 (17%) | 28 (-18;56) | 38 (-2;62) | 38/303 (13%) | 50 (28;65) | 63 (46;75) |
| Fully vaccinated household contacts | 164/1505 (11%) | 37/614 (6%) | 46 (22;63) | 46 (20;63) | 256/2070 (12%) | -16 (-44;6) | 40 (20;54) |

We previously found a higher VET to unvaccinated household contacts during the Alpha period (73% (95%CI 65-79))[1]. The secondary attack rate among unvaccinated contacts (22%) is also lower compared to the Alpha period (31%). Possibly, this is a result of increased prevalence of infection-induced immunity. In the beginning of August, around 20% of Dutch blood donors had infection-induced immunity [6]. Also, a larger share of index cases were of a younger age (below 30) compared to our Alpha period analysis, and SARs were lower for younger index cases (Supplementary Table S1). Our data does not contain information about negative tests. Therefore it is uncertain whether contacts tested negative or did not test at all. Even though both vaccinated and unvaccinated household contacts are advised to test at day 5 and in case of symptoms, we cannot preclude the possibility that testing rates among household contacts became lower, leading to an underestimation of the SAR. Differences in testing behavior between contacts of vaccinated and unvaccinated index cases could bias our VET estimates. During the study period, most Dutch adults had had the opportunity to receive vaccination, the coverage for 12 to 17-year-olds was still increasing during this period. The current vaccinated and unvaccinated populations are likely different in multiple aspects, such as risk behavior, willingness to test and adherence to quarantine. These aspects might bias out VET estimates in both directions: while the perceived risk of infection might be lower in vaccinated people due to their vaccination status, the perceived risk of infection among current unvaccinated populations could also be low due to personal beliefs. A lower risk perception for both groups may have resulted in decreased testing rates. Daily testing numbers averaged around 60,000 in spring 2021 whilst in August and September this averaged around 20,000, which is also likely influenced by the increasing use of at-home rapid antigen tests [7]. Further, as Table 2 shows, vaccinated and unvaccinated people are highly clustered within households. This reduces the power of our analysis.

It is known from literature that the Delta variant is more transmissible than Alpha, therefore a reduced VET for Delta compared to Alpha is not unexpected [8]. A recent study from the United Kingdom reported reduced transmission for vaccinated index cases, with aOR estimates in line with our VET estimates for both Alpha and Delta [9]. This study found that VET waned over time since vaccination of the index case. We explored whether such waning is also visible in our data (Supplementary Table S2). VET estimates are indeed lower when the index reached full vaccination status 60 or more days ago. However, our data does not allow detailed analysis of VET waning due to small numbers and strong correlation with age and time since vaccination of the household contacts. If VET indeed wanes over time, the lower VET for Delta compared to Alpha might be (partly) due to longer time since vaccination rather than the variant itself.

Our results indicate that vaccination confers protection against onward transmission from vaccinated index cases, albeit somewhat less for Delta than for Alpha. Vaccine effectiveness against transmission to unvaccinated household contacts is stronger than to vaccinated household contacts, with the latter already largely protected from infection, and especially from severe disease, by their own vaccine-induced immunity, but differences in risk behavior may also play a role. Possible waning of vaccine effectiveness against infection and against onward transmission could result in increases in SARS-CoV-2 circulation among populations with high vaccine coverage. As full vaccination remains highly effective in preventing severe disease, also for Delta, a high vaccination coverage remains the key to control the COVID-19 pandemic [10].

## Data Availability

The data are not publicly available.

## Acknowledgments

The authors would like to thank all source and contact tracing personnel at the 25 Municipal Health Services (GGDen) who have been invaluable for control and surveillance of the COVID-19 epidemic in the Netherlands, and who collected these important data.

## Members of the RIVM COVID-19 surveillance and epidemiology team

Agnetha Hofhuis, Anne Teirlinck, Alies van Lier, Bronke Boudewijns, Miek de Dreu, Anne-Wil Valk, Femke Jongenotter, Carolien Verstraten, Gert Broekhaar, Guido Willekens, Irene Veldhuijzen, Jan Polman, Jan van de Kassteele, Jeroen Alblas, Janneke van Heereveld, Janneke Heijne, Kirsten Bulsink, Lieke Wielders, Liselotte van Asten, Liz Jenniskens, Loes Soetens, Maarten Mulder, Maarten Schipper, Marit de Lange, Naomi Smorenburg, Nienke Neppelenbroek, Patrick van den Berg, Priscila de Oliveira Bressane Lima, Rolina van Gaalen, Sara Wijburg, Shahabeh Abbas Zadeh Siméon de Bruijn, Senna van Iersel, Stijn Andeweg, Sjoerd Wierenga, Susan Lanooij, Sylvia Keijser, Tara Smit, Don Klinkenberg, Jantien Backer, Pieter de Boer, Scott McDonald, Amber Maxwell, Annabel Niessen, Brechje de Gier, Danytza Berry, Daphne van Wees, Dimphey van Meijeren, Eric R.A. Vos, Frederika Dijkstra, Jeanet Kemmeren, Kylie Ainslie, Marit Middeldorp, Marjolein Kooijman, Mirjam Knol, Timor Faber, Albert Jan van Hoek, Eveline Geubbels, Birgit van Benthem, Hester de Melker, Jacco Wallinga, Rianne van Gageldonk-Lafeber, Susan Hahné, Susan van den Hof

