## Supplementary for "Vaccine effectiveness against SARS-CoV-2 transmission to household contacts during dominance of Delta variant (B.1.617.2), August-September 2021, the Netherlands"

Table S1. Secondary attack rates (SAR) for unvaccinated and fully vaccinated index cases, by age group of the index case.

| Age group index | Unvaccinated index - infected contacts / all contacts (SAR) | Fully vaccinated index - infected contacts / all contacts (SAR) |
|---|---|---|
| Unvaccinated household contacts | | |
| All ages | 547/2517 (22%) | 38/303 (13%) |
| 12-17 | 139/967 (14%) | 2/7 (29%) |
| 18-29 | 140/719 (19%) | 6/57 (11%) |
| 30-49 | 187/618 (30%) | 15/158 (9%) |
| 50-74 | 76/202 (38%) | 12/77 (16%) |
| 75+ | 5/11 (45%) | 3/4 (75%) |
| Fully vaccinated household contacts | | |
| All ages | 164/1505 (11%) | 256/2070 (12%) |
| 12-17 | 88/851 (10%) | 2/86 (2%) |
| 18-29 | 37/483 (8%) | 46/838 (5%) |
| 30-49 | 31/135 (23%) | 45/388 (12%) |
| 50-74 | 6/33 (18%) | 141/680 (21%) |
| 75+ | 2/3 (67%) | 22/78 (28%) |

Table S2. Secondary attack rate of SARS-CoV-2 and VET adjusted for time since full vaccination of the contact (< or >= 60 days, only in analysis of fully vaccinated contacts), age group of the index case and contact and week of notification date of the index case, stratified by time since full vaccination of the index case.

| Analysis | Unvaccinated index - infected contacts / all contacts (SAR) | Index fully vaccinated < 60 days ago - infected contacts / all contacts (SAR) | Index fully vaccinated < 60 days ago - adjusted VET (%) (95% CI) | Index fully vaccinated >= 60 days ago - infected contacts / all contacts (SAR) | Index fully vaccinated >= 60 days ago - adjusted VET (%)(95% CI) |
|---|---|---|---|---|---|
| Unvaccinated household contacts | 547/2517 (22%) | 24/209 (11%) | 67 (47;79) | 14/94 (15%) | 55 (19;76) |
| Fully vaccinated household contacts | 164/1505 (11%) | 99/1278 (8%) | 57 (40;69) | 157/792 (20%) | 28 (-4;50) |
